# Favipiravir In Adults with Moderate to Severe COVID-19: A Phase 3 Multicentre, Randomized, Double-Blinded, Placebo-Controlled Trial

**DOI:** 10.1101/2021.11.08.21265884

**Authors:** Srinivas Shenoy, Sagar Munjal, Sarah Al Youha, Mohammad Alghounaim, Sulaiman Almazeedi, Yousef Alshamali, Richard H Kaszynski, Salman Al-Sabah, Kuwait Clinical Trial Group

## Abstract

**Aim:** To assess the efficacy and safety of favipiravir in adults with moderate to severe coronavirus disease 2019 (COVID-19).

**Methods:** In this randomized, double-blind, multicenter, phase 3 trial, adults (21-80 years) with real-time reverse transcriptase polymerase chain reaction (rRT-PCR) confirmed SARS-CoV-2 infection and presenting with moderate to severe COVID-19 and requiring hospitalization were randomized 1:1 to oral favipiravir (day 1: 1800 mg BID and days 2-10: 800 mg BID) (FPV) plus standard supportive care (SoC) versus placebo plus SoC (placebo). The primary endpoint was time to resolution of hypoxia.

**Results:** In total, 353 patients were randomized to receive either FPV or placebo (175 and 178 in the FPV and placebo groups, respectively). Overall, 76% of the patients (240/315, 78% in FPV vs. 75% in placebo group) reached resolution of hypoxia on or before day 28. The median time to resolution of hypoxia was 7 days in the FPV group and 8 days in the placebo group. Treatment effect was not significant [Hazard ratio (HR) (95% CI): 0.991 (0.767, 1.280) (*p*=0.94)].

Patients in the lower NEWS-2 clinical risk subgroup were more likely to achieve shorter time to resolution of hypoxia with the median time to resolution of hypoxia of 6 days in FPV and 7 days in placebo group [HR (95% CI): 1.21 (0.847, 1.731) (*p*=0.29)]; shorter time to hospital discharge with a median time to discharge of 8 and 10 days in the FPV and placebo group, respectively [HR (95% CI): 1.47 (1.081, 1.997) (*p=*0.014)]; and shorter time to improvement by 1-point improvement over baseline in WHO 10-point clinical status score with the median time to improvement by 1-point from baseline of 6 and 7 days in the FPV and placebo group, respectively [HR (95% CI): 1.16 (0.830, 1.624) (*p=*0.38)] than higher NEWS-2 clinical risk subgroup.

Treatment emergent adverse event (TEAEs) were experienced by 62/334 (19%) patients [35/168 (21%) patients in FPV and 27/166 (16%) in placebo group]. Hyperuricaemia/increased blood uric acid was reported in 9 (3%)/2 (1%) patients [8 (5%)/1(1%) patients in FPV and 1 (1%)/1(1%) in placebo group], which were of mild intensity and transient. Overall, 36 serious adverse events (SAEs) were reported, 20 in FPV and 16 in placebo group.

**Conclusion:** The trial did not find favipiravir to be effective in moderate to severe, hospitalized COVID-19 patients; favourable clinical trends were observed in patients with lower NEWS-2 risk when early administration of favipiravir could be achieved.

## INTRODUCTION

The coronavirus disease 2019 (COVID-19) pandemic has highlighted the need for safe and effective anti-viral treatments. This is particularly true for ‘high risk’ patient groups, such as the elderly and patients with underlying co-morbidities, who are more likely to experience poor outcomes from COVID-19[1,2]. Despite large scale vaccination efforts worldwide, a significant portion of the global population remain unvaccinated due to supply shortages and general vaccine hesitancy[3]. In addition, the emergence of variants such as Delta (B.1.617.2) continue to pose a threat owing to higher transmissibility, disease severity and ability to evade immune response elicited by severe acute respiratory syndrome coronavirus 2 (SARS-CoV-2) vaccination[4].

A wide array of therapeutic agents are currently undergoing clinical trials including many repurposed antiviral drugs[5]. Favipiravir, an antiviral drug approved in Japan in 2014 to treat novel or re-emerging influenza virus infections, was evaluated as a potential therapeutic option during the COVID-19 pandemic. Favipiravir has also been employed in the treatment of Ebola during the 2014 epidemic and additional indications include severe fever with thrombocytopenia virus, rabies, Lassa fever, Jamestown Canyon virus, and norovirus[6–8]. The convenience of being an orally administered low-cost therapeutic, along with its inherent thermostability make the drug an appealing candidate for COVID-19 treatment, especially in low-income countries with limited access to novel vaccines and therapeutics.

Favipiravir (T-705; 6-fluoro-3-hydroxy-2-pyrazinecarboxamide) is an anti-viral agent that selectively and potently inhibits the RNA-dependent RNA polymerase (RdRp) of RNA viruses.[6] An *in-vitro* study demonstrated that favipiravir effectively inhibits the SARS CoV-2 infection in Vero E6 cells (ATCC-1586). An optimal dosing regimen was derived based on the *in-vitro* study findings and prior clinical trial experience[9,10].

Though several anecdotal reports and observational studies have reported a positive trend for Favipiravir in hospitalized moderate to severe COVID-19 patients, none of these studies were double-blinded or adequately powered. The present study is a double-blind, placebo-controlled, phase 3 using Favipiravir administered in 1800/800 mg doses with a 10-day dosing regimen in hospitalised COVID-19 patients with moderate to severe disease. Earlier studies have established safety profiles with the proposed regimen for up to 22 days[8–12].

## METHODS

This was a prospective, interventional, multicentre, randomised (1:1), double-blind, placebo-controlled, parallel design phase 3 trial to evaluate the efficacy, safety and tolerability of favipiravir (FPV) with supportive care (SoC) in comparison to placebo with SoC in the acute treatment of hospitalized patients who tested positive for SARS-CoV-2 and presented with moderate to severe COVID-19. The study was conducted at three hospitals in Kuwait [Jaber Al Ahmad Al Jaber Al Sabah Hospital, Mishref Field Hospital (both of which are designated COVID-19 centers in Kuwait) and Farwaniya Hospital] in the period between 22 August 2020 and 27 January 2021, after obtaining ethical approval from the Ministry of Health (MoH), Kuwait. The study was registered on clinicaltrials.gov under the registration number: NCT04529499.

### Participating Patients

The study included patients of either sex, aged between 21 (age of giving informed consent in Kuwait) and 80 years (both inclusive) who had tested positive for SARS-CoV-2 by real-time Reverse Transcriptase Polymerase Chain Reaction (rRT-PCR) assay on a nasopharyngeal or oropharyngeal swab, were clinically assessed to have moderate or severe COVID-19 and were hospitalized for management. Patients that agreed to participate in the study signed an informed consent statement. Critically ill patients and patients who had first onset of symptoms/signs suggestive of COVID-19 illness >10 days before randomization were excluded from the study. Other main exclusion criteria were patients who had allergy or contraindication to the drug, pregnant and lactating mothers, and patients with congestive cardiac failure, moderate to severe hepatic dysfunction or renal failure, alanine aminotransferase (ALT) and/or aspartate aminotransferase (AST) levels > 5 times upper limit of normal (ULN) at screening evaluation, serum uric acid higher than the ULN at screening evaluation, those with history of gout or were on current treatment for gout.

### Trial Design

The study was conducted in two parts: Stage I – Main study: Days 1 to 28 [telephonic follow up of patients was performed after discharge from hospital (on Days 10, 14, 21 and 28, as applicable)]; and Stage II – Extended follow up: Days 29 to 60 (telephonic follow up assessments were performed on Day 42 and 60, in case of patients discharged from hospital in Stage I). The treatment duration was ten consecutive days. The dose of favipiravir (FPV) selected for the study was a loading dose of 1800 mg BID (200 mg, 9 tablets BID) on Day 1 followed by 800 mg BID (200 mg, 4 tablets BID) for the next nine days. Matching no. of placebo tablets were used for patients randomized to control group.

Screening investigations, including clinical laboratory evaluations (hematology, serum biochemistry, urinalysis, serum pregnancy test), chest X-ray, ECG and rRT-PCR assay for detection of SARS-CoV-2 RNA, were performed within 72 hours before randomization/baseline visit (Day 1) for confirmation of eligibility. During the study, clinical status was evaluated by the WHO 10-point ordinal scale, National Early Warning Score 2 (NEWS-2) score and COVID-19 associated symptom severity (i.e., presence or absence, and if present, the severity of individual COVID-19 related symptoms). Vital signs and physical examination were performed every day (until discharge or Day 28, whichever was earlier). Chest X-rays were performed on days 4, 7, 10, 14, 28 or discharge or study discontinuation, whichever is earlier. A respiratory tract sample (either a nasopharyngeal swab, oropharyngeal swab, nasal aspirate, tracheobronchial aspirate or bronchial lavage) was collected from the patient and sent for detection (qualitative) of SARS-CoV-2 RNA by rRT-PCR assay on days 5, 10 or upon discharge. Safety assessments included serum pregnancy tests for female patients of child-bearing potential, 12-Lead ECG, clinical laboratory evaluations (hematology, serum biochemistry, urinalysis). Treatment-emergent AEs and concomitant medication were recorded.

NEWS-2 score was used as an aid to clinical assessment and patients were categorized for clinical risk using baseline NEWS-2 scores, as ‘low’ (aggregate score of 0-4), ‘low-medium’ (Score of 3 in any individual parameter), ‘medium’ (aggregate score 5–6) and ‘high’ (aggregate score 7 or more)[13]. [NEWS-2 scale presented in Supplementary Data] For discussion of the results of subgroup analyses, patients have been categorized into two ‘clinical risk’ subgroups based on baseline NEWS-2 scores: a ‘lower’ risk subgroup (which consists of patients in ‘low’ or ‘low-medium’ baseline NEWS-2 clinical risk categories) and ‘higher’ risk subgroup (which consists of patients in ‘medium’ or ‘high’ risk NEWS-2 categories) in this report.

Compliance to study treatment was assessed daily when the patients were hospitalized. For patients who were discharged before the end of treatment period (Day 10), the compliance was assessed based on the details in the investigational medicinal product (IMP) accountability log/patient diary and the empty blister packs and/unused IMP.

The study’s primary objective was to evaluate the efficacy of oral FPV in improving the time to resolution of hypoxia. The study’s secondary objectives were to evaluate the efficacy of oral FPV on clinical and virological outcomes in moderate to severe COVID-19 patients and to assess the safety and tolerability of FPV[14].

Time to resolution of hypoxia, the primary endpoint of this study, was considered to have been met when the patient had attained a score of 4 or lower on the WHO 10-point ordinal scale of clinical status (and either had a score ≤ 4 on consecutive assessments over the next 5 days if patient continued to remain in hospital, or the patient was discharged before five consecutive assessments after reaching a score of 4 and the patient had survived and had not been readmitted to hospital for COVID-19 management till Day 28).

### Statistical Methods

Inequality testing of the hazard ratio using the Cox proportional hazards (CPH) regression model with 371 subjects in Favipiravir group and 371 subjects in Placebo group achieved 80% power at the 0.05 significance level for an actual hazard ratio of 1.25 assuming the hazard ratio is 1 under the null hypothesis and that the total number of events achieved is 631. Based on the above assumption and considering the dropout rate to be 5% overall, 390 patients were to be randomized in each treatment group. Sample size re-estimation was provisioned at interim analysis.

The continuous endpoints were summarized using descriptive statistics and categorical data were summarized using counts and percentages. Unadjusted between-treatment comparisons were made using Fisher’s exact test (binary variables), Chi-square test (multi-categorical variables), Student’s t-test (continuous variables), and Kaplan-Meier estimates / log-rank test (time to event variables). The primary endpoint, time to resolution of hypoxia, was compared between the treatment groups using the Cox proportional hazards model with age and gender as covariates. Two-sided p-values of less than 0.05 were considered statistically significant. The SAS^®^ package, Version 9.4, was used for statistical evaluation.

A data monitoring committee (DMC), consisting of 3 members specializing in critical care and a statistician, assessed emerging safety and efficacy data from the trial and made recommendations on study continuation after a single pre-specified interim analysis.

## RESULTS

The scheduled interim analysis was performed on the data collected for the first 349 randomized patients (patients who were enrolled before the cutoff date of 31 December 2020), including 173 in the FPV and 176 in the placebo group, and was submitted for DMC review. The DMC recommended study termination due to futility (lack of evidence for efficacy) in the pre-specified primary endpoint, time to resolution of hypoxia, and key secondary endpoints: time to hospital discharge and mortality. In all three analyses, the estimated conditional power was very low (less than 10%).

A total of 353 patients (ITT population, which consisted of all randomized patients; including four patients who were in the treatment phase and not included for the interim analysis) were randomized to 2 treatment groups, 175 in FPV + SoC and 178 in placebo + SoC group. Among the 353 patients randomized, 190 (53.8%) patients completed Stage I of the study, with both the groups having 95 (54.3% in FPV and 53.4% in placebo) patients each. The reasons for early termination from study included death due to COVID-19 or COVID-19 associated complication [25 (32.9%)], withdrawal of consent [19 (25.0%)], adverse events [8 (10.5%)], lost to follow-up [2 (2.6%)] and other reasons [18 (23.7%)].

**Figure 1:**
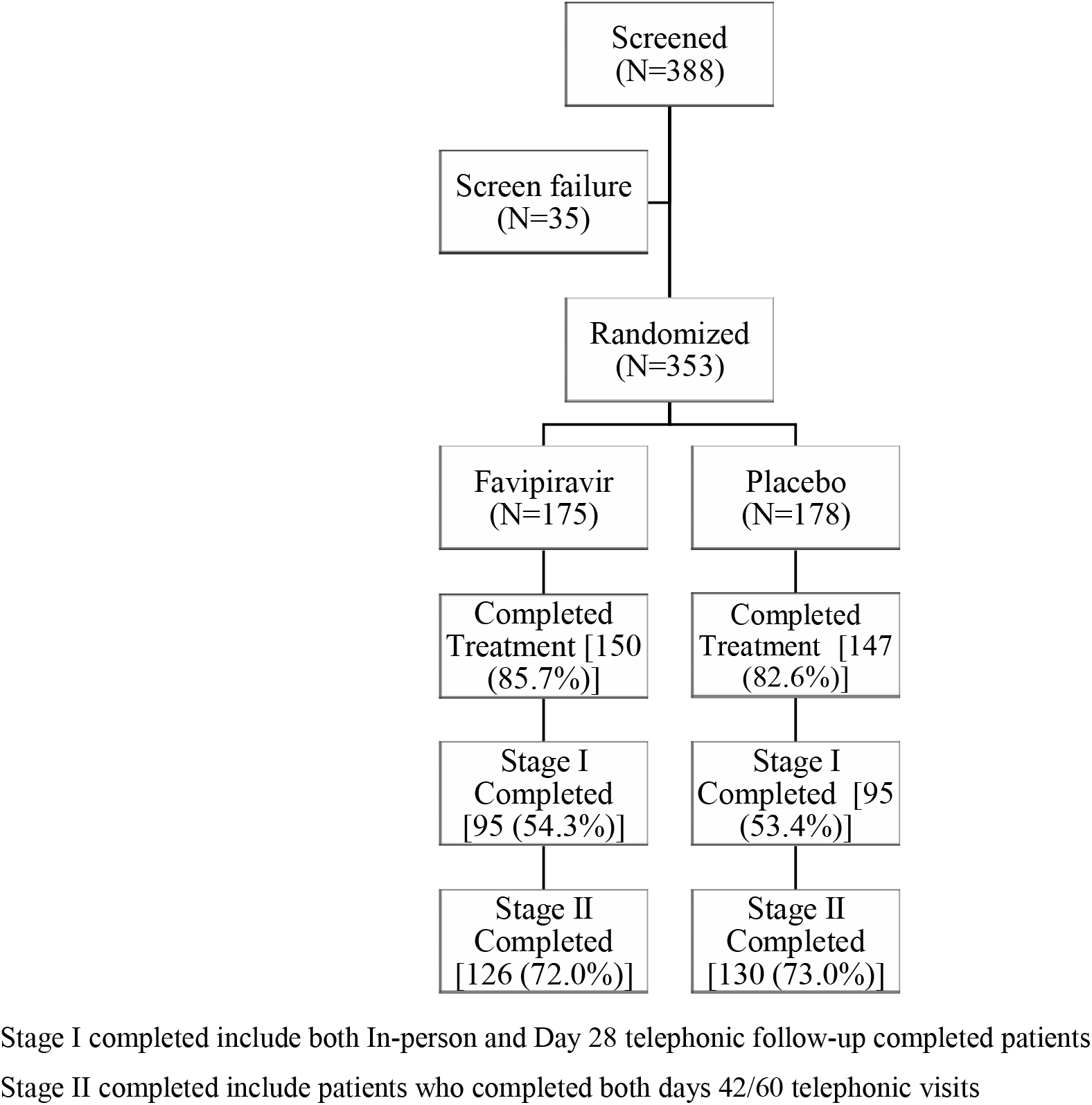
CONSORT Diagram of Patient Disposition.

The two treatment groups were balanced with respect to demographic characteristics and COVID-19 disease severity and symptoms at baseline. The mean (Standard deviation, SD) age of the patients randomized in the study was 51.9 (±12.5) years. There were 144 (40.8%) patients over the age of 50 years and the majority of the patients were male [238 (67.4%)]. At baseline, the median number of days since onset of first symptom(s) associated with COVID-19 was 7.0 (IQR 5.0 - 7.0) in both groups.

**Table 1:** Patient Demographic Characteristics at Screening and COVID-19 Severity and Symptoms at Baseline, ITT population.

| Variable | Statistics/<br>Response<br>Category | Total<br>(N=353) | By Treatment Group |  | P-Value* |
| --- | --- | --- | --- | --- | --- |
|  |  |  | Favipiravir<br>(N=175) | Placebo<br>(N=178) |  |
| Gender | Male | 238 (67.4%) | 118 (67.4%) | 120 (67.4%) | 1.00 |
|  | Female | 115 (32.6%) | 57 (32.6%) | 58 (32.6%) |  |
| Age group (years) | <50 | 144 (40.8%) | 70 (40.0%) | 74 (41.6%) | 0.83 |
|  | 50+ | 209 (59.2%) | 105 (60.0%) | 104 (58.4%) |  |
| Clinical Status by WHO 10-Point Ordinal Scale | 4= Hospitalized, no oxygen therapy | 38 (10.8%) | 18 (10.3%) | 20 (11.2%) | 0.78 |
|  | 5= Hospitalized, on oxygen | 312 (88.4%) | 156 (89.1%) | 156 (87.6%) |  |
|  | 6= Hospitalized, Oxygen by NIV or high flow | 2 (0.6%) | 1 (0.6%) | 1 (0.6%) |  |
|  | 7= MV, p/f>150 or s/f >200 | 1 (0.3%) | 0 (0.0%) | 1 (0.6%) |  |
| NEWS Clinical Risk | Low (Score: 0-4) | 182 (51.6%) | 86 (49.1%) | 96 (53.9%) | 0.61 |
|  | Low-Medium (Score of 3 in any individual parameter) | 1 (0.3%) | 0 (0.0%) | 1 (0.6%) |  |
|  | Medium (Score: 5-6) | 143 (40.5%) | 74 (42.3%) | 69 (38.8%) |  |
|  | High (Score: ≥7) | 24 (6.8%) | 14 (8.0%) | 10 (5.6%) |  |
|  | Data Missing | 3 (0.8%) | 1 (0.6%) | 2 (1.1%) |  |
| Randomization Severity Strata** | Moderate | 256 (72.5%) | 127 (72.6%) | 129 (72.5%) | 0.98 |
|  | Severe | 97 (27.5%) | 48 (27.4%) | 49 (27.5%) |  |
| SpO<sub>2</sub> on Room Air or SpO<sub>2</sub> / FiO<sub>2</sub> ratio on Supplemental Oxygen | Moderate | 153 (43.3%) | 78 (44.6%) | 75 (42.1%) | 0.83 |
|  | Severe | 195 (55.2%) | 95 (54.3%) | 100 (56.2%) |  |
|  | Data Missing | 5 (1.4%) | 2 (1.1%) | 3 (1.7%) |  |
| Number of Days Since First Onset of COVID-19 Symptoms | Mean (SD) | 6.3 (1.8) | 6.3 (1.8) | 6.2 (1.7) | 0.57 |
|  | Median | 7.0 | 7.0 | 7.0 |  |
|  | IQR (Q1, Q3) | 2.0 (5.0, 7.0) | 2.0 (5.0, 7.0) | 2.0 (5.0, 7.0) |  |

N: Total number of patients in the specified treatment group; IQR: Interquartile IQR=Q3 – Q1; NIV: Non-invasive ventilation; MV: mechanically ventilated; p/f ratio: PaO<sub>2</sub>/FiO<sub>2</sub> Ratio; s/f ratio: SpO<sub>2</sub> to FiO<sub>2</sub> ratio; SD: Standard deviation.
Percentages are based on the total number of patients randomized for the study.
\* Continuous variable (Student's t-test), binary variable (Fisher's exact test), multi-category variable (Chi-square test)
\*\* Based on the FDA Guidance document COVID-19: Developing Drugs and Biological Products for Treatment or Prevention - Guidance for Industry Final Document dated May 2020

At randomization, on the WHO 10-point ordinal scale of clinical status, 38 (10.8%) [18 (10.3%) in FPV and 20 (11.2%) in placebo] had a score of 4 (hospitalized: no oxygen therapy) and 312 (88.4%) patients [156 in both groups; 89.1% in FPV and 87.6% in placebo] had a score of 5 (hospitalized, on oxygen) two patients (0.6%) had a score of 6 (hospitalized, oxygen by non-invasive ventilation or high flow) and one patient from the placebo group had a score of 7 (mechanically ventilated, PaO2/FiO2>150 or SpO2/FiO2 >200).

In the current study, 183 (51.8%) patients were classified as ‘lower’ clinical risk at baseline, which included ‘low’ clinical risk in 182 (51.6%) patients [86 (49.1%) in FPV and 96 (53.9%) in placebo] and ‘low-medium’ clinical risk in 1 (0.6%) patient in placebo group. The baseline clinical risk was classified as ‘higher’ in 167 (47.3%) patients, which included ‘medium’ clinical risk in 143 (40.5%) patients [74 (42.3%) in FPV and 69 (38.8%) in placebo] and ‘high’ in 24 (6.8%) patients [14 (8.0%) patients in FPV and 10 (5.6%) in placebo]}. Baseline NEWS-2 scores were missing for 3 (0.8%) [1(0.6%) in FPV and 2 (1.1%) in placebo group] patients.

Overall, a mean (±SD) compliance of 81.0 % (±22.3), with 79.8% (±22.3) in FPV and 82.1% (±22.3) in placebo group was observed during the study.

### Efficacy

#### Primary endpoint analysis

The population for the primary endpoint analysis included patients with baseline WHO 10-point ordinal scale of clinical status score of 5 or higher and consisted of 315 patients overall, with 157 (89.7%) in FPV and 158 (88.8%) in placebo group. The population for the primary endpoint analysis was 89.2% of the ITT population included in the study.

##### Time to Resolution of Hypoxia

The median time to resolution of hypoxia was 7 days in the FPV and 8 days in placebo group, as per Kaplan-Meier estimates. Treatment effect was not significant [HR (95% CI): 0.99 (0.767, 1.280) (*p*=0.94)].

In a *post-hoc* subgroup analysis, a weak trend in favor of FPV treatment was observed in patients classified under ‘lower’ clinical risk subgroup with the median time to resolution of hypoxia of 6 days in FPV and 7 days in placebo group [Log-rank test: *p=*0.26; HR (95% CI): 1.21 (0.847, 1.731) (*p*=0.29)]. In the ‘higher’ clinical risk subgroup, the median time to resolution of hypoxia was 8 days in both groups [Log-rank test: *p=*0.83; HR (95% CI): 0.96 (0.672, 1.381) (*p*=0.84)].

**Figure 2:**
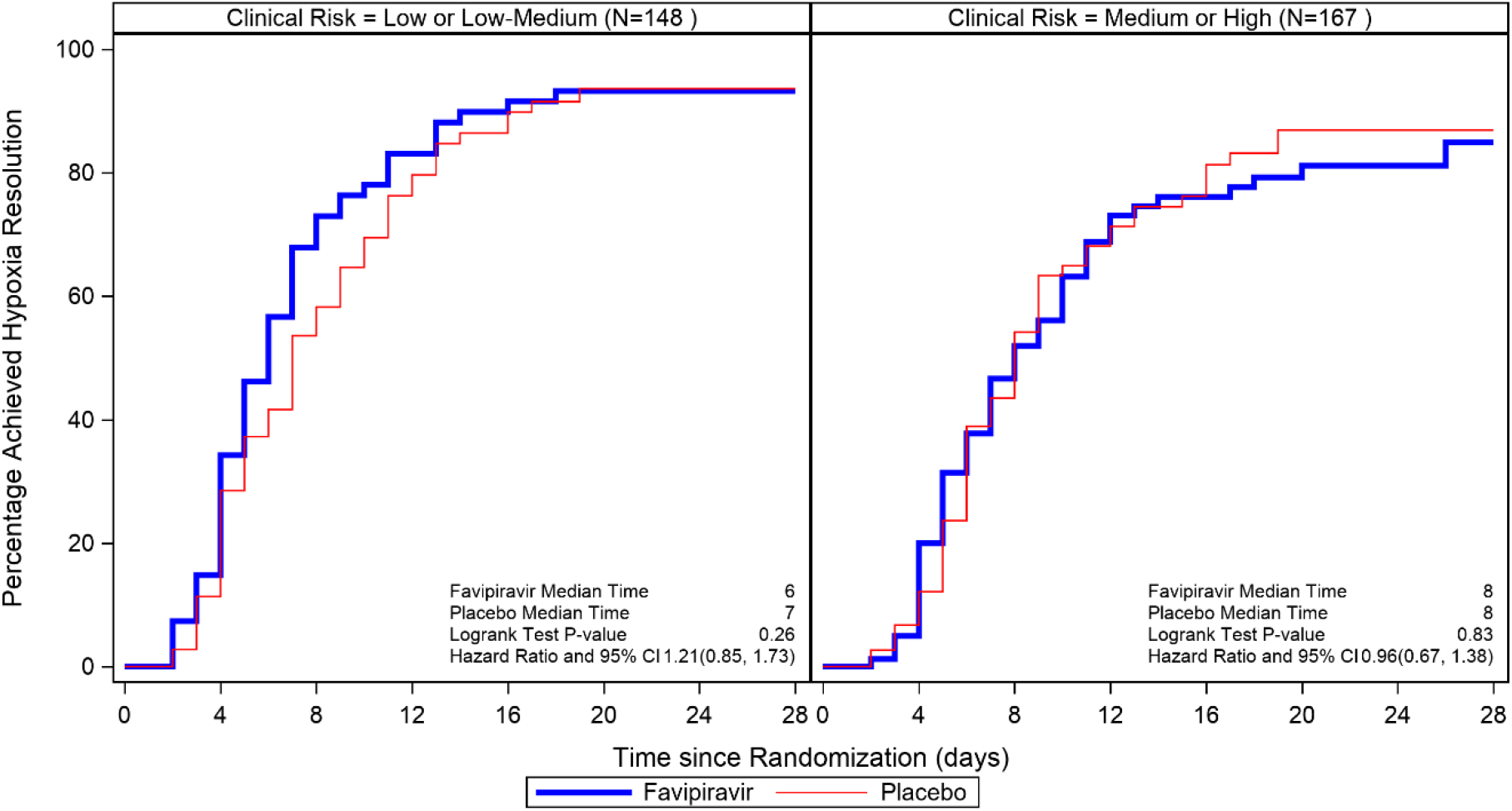
Kaplan-Meier Estimates of Time to Resolution of Hypoxia by NEWS-2 Clinical Risk, ITT Population with Baseline WHO 10-Point Clinical Status Score > 4 (N=315)

**Table 2:** Analysis of Time to Event Endpoints, ITT Population.

| Parameters/Variables | Favipiravir<br>(N=175) | Placebo<br>(N=178) |
| --- | --- | --- |
| ***Time to Resolution of Hypoxia (Primary endpoint)*** |  |  |
| No of Patients* | 157 | 158 |
| No of Events [Time frame: Up to 28 days]<br>(% Reached Resolution Endpoint) | 122 (77.7%) | 118 (74.7%) |
| Time to event, median days | 7 | 8 |
| Hazard Ratio (95% CI) | 0.99 (0.767, 1.280) | - |
| P-value<sup>§</sup> | 0.94 | - |
| ***Time to Hospital Discharge by Treatment*** |  |  |
| No of Events (% Reached Discharge Endpoint) | 157 (89.7%) | 156 (87.6%) |
| Time to event, median days | 10 | 11 |
| Hazard Ratio (95% CI) | 1.06 (0.849, 1.326) | - |
| P-value<sup>§</sup> | 0.60 | - |
| ***Time to improvement by 1-point from baseline in WHO 10-point clinical status score*** |  |  |
| No of Events (% Reached Improve 1-Point Endpoint) | 129 (73.7%) | 129 (72.5%) |
| Time to event, median days | 7 | 8 |
| Hazard Ratio (95% CI) | 1.05 (0.820, 1.337) | - |
| P-value<sup>§</sup> | 0.71 | - |
| ***Time to improvement by 2-point from baseline in WHO 10-point clinical status score*** |  |  |
| No of Events (% Reached Improve 2-Point Endpoint) | 18 (10.3%) | 28 (15.7%) |
| Time to event, median days | - | - |
| Hazard Ratio (95% CI) | 0.67 (0.372, 1.216) | - |
| P-value<sup>§</sup> | 0.19 | - |

CI: Confidence Interval
\* Patients with Baseline WHO 10-Point Clinical Status Score > 4 (Hospitalized: no oxygen therapy) (N=315) were included in the Primary endpoint analysis (Time to Resolution of Hypoxia); 38 patients with score of 4 were not included.
§ From Cox proportional hazards model

#### Secondary endpoint Analysis

##### Time to hospital discharge

The median time to hospital discharge was 10 days in the FPV and 11 days in placebo group. No significant differences were observed between the FPV and placebo groups [HR (95% CI): 1.06 (0.849, 1.326) (*p=*0.60)].

In a *post-hoc* subgroup analysis, a significant difference was noted in time to hospital discharge in patients classified under the ‘lower’ clinical risk subgroup, with a median time to discharge of 8 and 10 days in the FPV and placebo group, respectively [Log-rank test: *p=*0.0062; HR (95% CI): 1.47 (1.081, 1.997) (*p=*0.014)]. In the ‘higher’ clinical risk subgroup, the median time to discharge was 11 days in both treatment groups [Log-rank test: *p=*0.59; HR (95% CI): 0.92 (0.665, 1.272) (*p=*0.61)].

**Figure 3:**
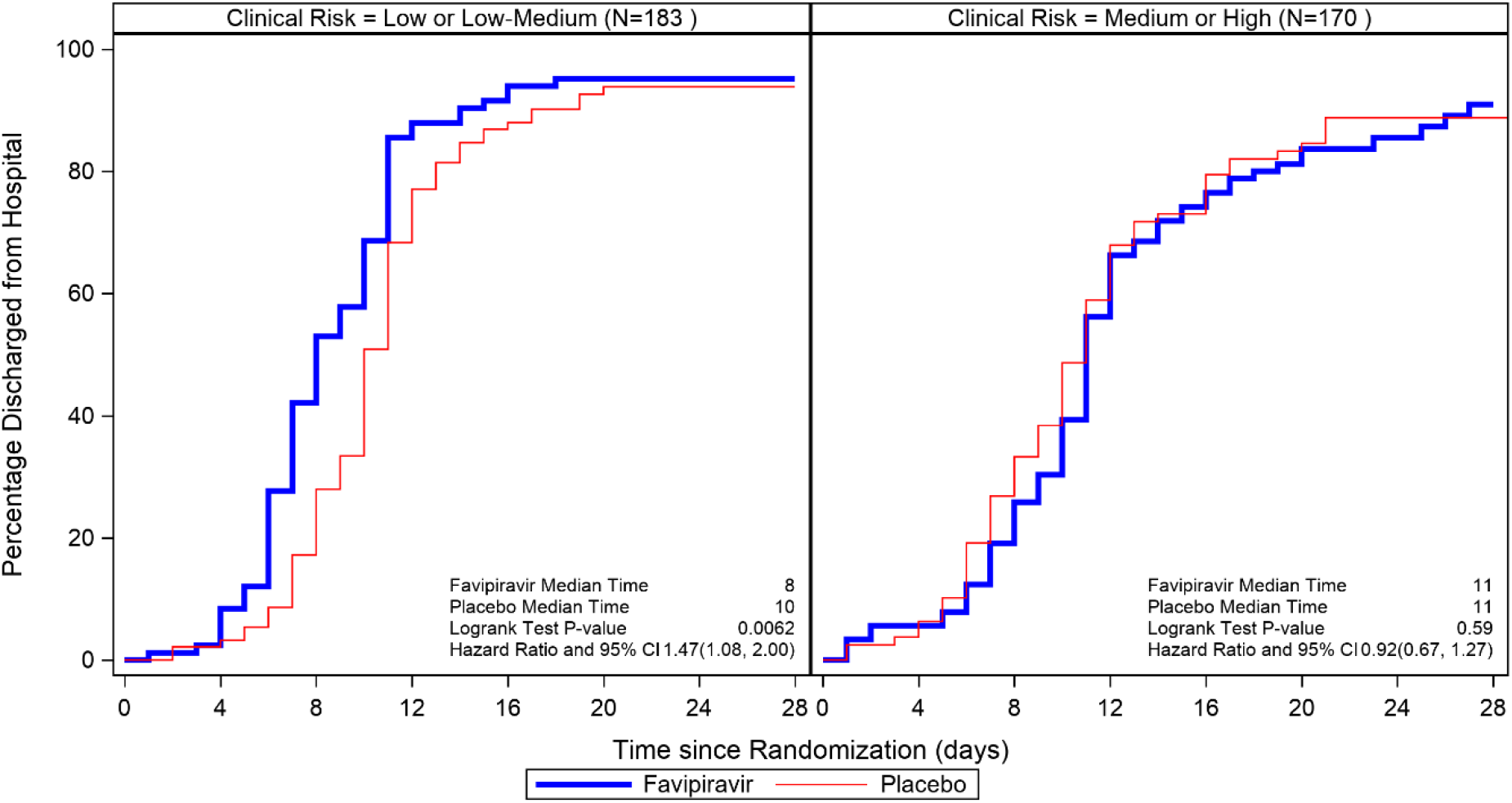
*Post-hoc* Analysis: Kaplan-Meier Estimates of Time to Hospital Discharge by Baseline NEWS-2 Clinical Risk Categories, ITT Population (N=353)

##### Time to improvement by 1 and by 2 points over baseline in WHO 10-point clinical status score

The median time to 1-point improvement over baseline in WHO 10-point clinical status score was 7 days in the FPV and 8 in placebo group, as per Kaplan-Meier estimates. Treatment effect was not significant [HR (95% CI): 1.05 (0.820, 1.337) (*p=*0.71)].

In a *post-hoc* subgroup analysis, a weak numerical trend in favour of the FPV treatment was observed in the patients classified under ‘lower’ clinical risk subgroup, with the median time to improvement by 1-point from baseline of 6 and 7 days in the FPV and placebo group, respectively [Log-rank test: *p=*0.35; HR (95% CI): 1.16 (0.830, 1.624) (*p=*0.38)]. In the ‘higher’ clinical risk subgroup, the median time to improvement by 1-point from baseline was 8 days in both the treatment groups [Log-rank test: *p=*0.87; CPH model HR (95% CI): 0.97 (0.679, 1.390) (*p=*0.87)].

**Figure 4:**
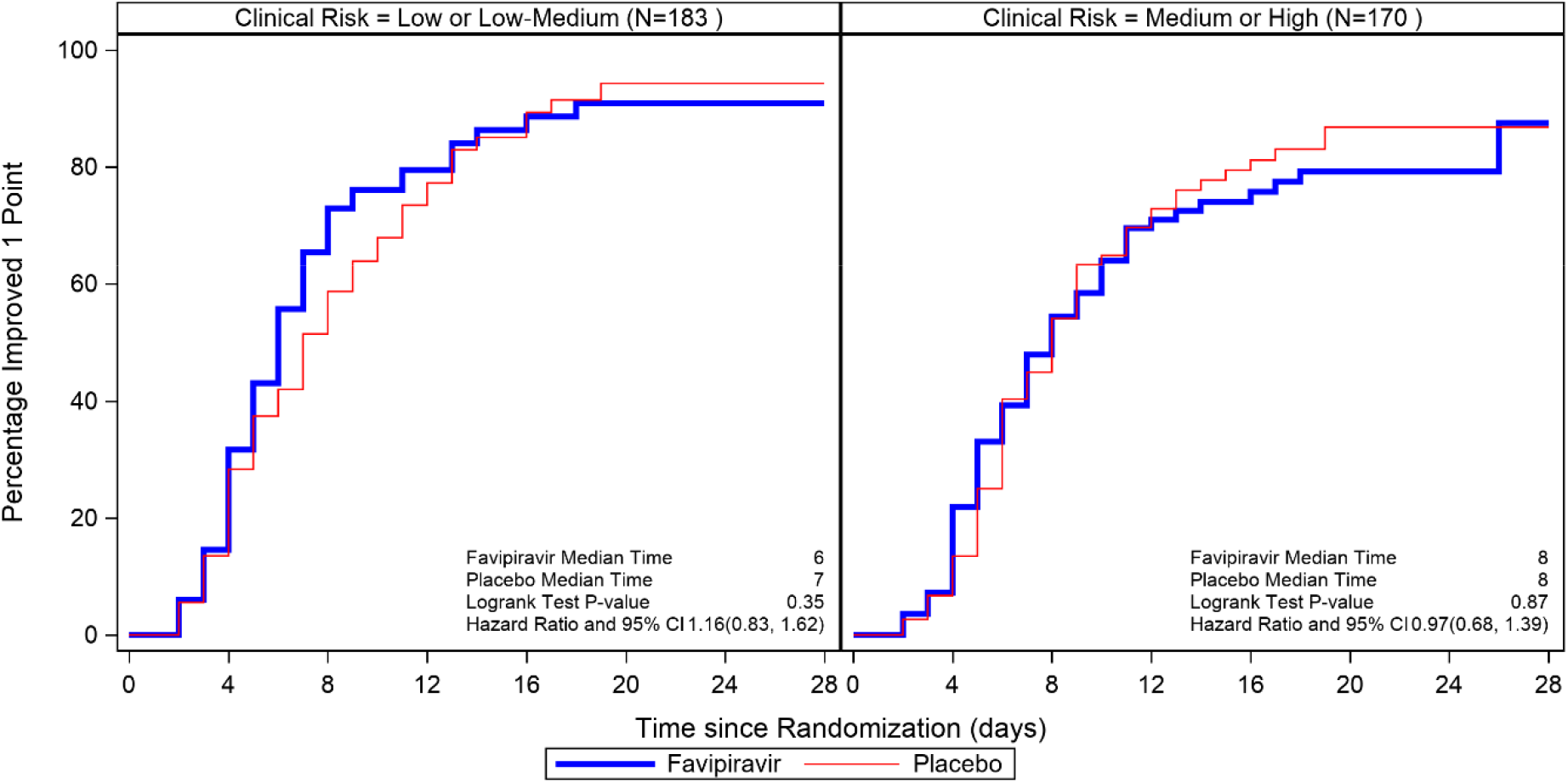
*Post-hoc* Analysis: Kaplan-Meier Estimates of Time to Improve 1 Point in WHO 10-Point Clinical Status Score from Baseline by Baseline NEWS-2 Clinical Risk Categories, ITT Population (N=353)

**Table 3:** Summary of Subgroup Analysis of Time to Event Endpoints By Baseline NEWS-2 Clinical Risk Categories.

| NEWS-2 Clinical risk category* | Lower risk |  | Higher risk |  |
| --- | --- | --- | --- | --- |
| Parameters/Variables | Favipiravir<br>(N=86) | Placebo<br>(N=97) | Favipiravir<br>(N=89) | Placebo<br>(N=81) |
| ***Time to Resolution of Hypoxia (Primary endpoint)*** |  |  |  |  |
| No of Patients at clinical risk category ** | 70 | 78 | 87 | 80 |
| No of Events [Time frame: Up to 28 days]<br>(% Reached Resolution Endpoint) | 60 (85.7%) | 61 (78.2%) | 62 (71.3%) | 57 (71.3%) |
| Time to event, median days | 6 | 7 | 8 | 8 |
| Hazard Ratio (95% CI) | 1.21 (0.847, 1.731) | - | 0.96 (0.672, 1.381) | - |
| P-value <sup>§</sup> | 0.29 |  | 0.84 |  |
| ***Time to Hospital Discharge by Treatment*** |  |  |  |  |
| No of Events (% Reached Discharge Endpoint) | 79 (91.9%) | 86 (88.7%) | 78 (87.6%) | 70 (86.4%) |
| Time to event, median days | 8 | 10 | 11 | 11 |
| Hazard Ratio (95% CI) | 1.47 (1.081, 1.997) | - | 0.92 (0.665, 1.272) | - |
| P-value<sup>§</sup> | 0.014 | - | 0.61 | - |
| ***Time to improvement by 1-point from baseline in WHO 10-point clinical status score*** |  |  |  |  |
| No of Events (% Reached Improve 1 Point Endpoint) | 66 (76.7%) | 72 (74.2%) | 63 (70.8%) | 57 (70.4%) |
| Time to event, median days | 6 | 7 | 8 | 8 |
| Hazard Ratio (95% CI) | 1.16 (0.830, 1.624) | - | 0.97 (0.679, 1.390) | - |
| P-value<sup>§</sup> | 0.38 |  | 0.87 |  |

CI: Confidence Interval
\* Lower risk category consisted of Low or Low-Medium risk NEWS-2 categories; Higher risk category consisted of Medium or High-risk NEWS-2 categories
\*\* Patients with Baseline WHO 10-Point Clinical Status Score > 4 (Hospitalized: no oxygen therapy) (N=315) were included in the Primary endpoint analysis (Time to Resolution of Hypoxia); 38 patients with score of 4 were not included.
<sup>§</sup> From Cox proportional hazards model

Only 13% of the patients (46/353, 18 (10%) in the FPV and 28 (16%) in placebo group) reached 2-point improvement endpoint. Therefore, the median time to improve by 2-points over baseline on WHO 10-point clinical status score could not be calculated in either treatment group. Most patients [312 (88.4%)] were enrolled in the study with a score of 5 and were discharged on attaining a score 4.

##### Proportion of patients who attained WHO 10-point clinical status score improvement by 1 and by 2 points

The proportion of patients who attained improvement by at least 1-point over baseline was numerically higher in the FPV group on Day 4 [42 (24.0%) compared to 31 (17.4%) in placebo group], Day 7 [62 (35.4%) in FPV compared to 60 (33.7%) in placebo group], and Day 28 or discharge [118 (67.4%) in FPV compared to 105 (59.0%) in placebo group]. The number of patients who attained a WHO 10-point clinical status score improvement by at least 2-points over baseline was numerically higher in the FPV group on Day 4 [3 (1.7%) compared to 1 (0.6%) in the placebo group]. [*Tables presented in Supplementary Data*]

##### Proportion of patients with disease progression [management in ICU, requirement of high flow nasal oxygen or non-invasive mechanical ventilation (NIMV) or invasive mechanical ventilation (IMV) until Day 28 or discharge from hospital (if discharge happened earlier]

The percentage of patients who required management in ICU until Day 28 or discharged from hospital were equally distributed in both the treatment groups [40 (11.3%) overall, 20 (11.4%) in FPV and 20 (11.2%) in the placebo group].

Among the 44 (12.5%) patients who required high flow nasal oxygen or NIMV, more patients belonged to the placebo group [26 (14.6%)] than the FPV group [18 (10.3%)].

Among the 30 (8.5%) patients who required IMV, more patients belonged to FPV group [17 (9.7%)] compared to placebo group [13 (7.3%)].

##### Summary of Deaths recorded in the study by Treatment

Of the 25 (7.1%) deaths reported during the study, 14 (8.0%) were reported in FPV and 11 (6.2%) in placebo group (*p=*0.54).

#### Safety

Treatment emergent adverse events (TEAEs) were experienced by 62/334 (19%) patients [35/168 (21%) and 27/166 (16%) patients in the FPV and placebo group, respectively]. As observed in previous studies, FPV was found to be associated with transient elevations in blood uric acid. Hyperuricaemia/increased blood uric acid was reported in 9 (3%)/2 (1%) patients [8 (5%)/1(1%) and 1 (1%)/1(1%) patients in the FPV and placebo groups, respectively]. All were of mild intensity and transient in nature. TEAEs associated with liver included increased hepatic enzymes observed in 11 (3%) patients [6 (4%) in the FPV and 5 (3%) in placebo group], hepatitis in 2 patients [one patient each in both treatment groups], alanine aminotransferase elevated in one patient in the FPV group and liver function test increased in one patient in the placebo group only.

**Table 4:** TEAEs Stratified by System Organ Class (>1%) and Preferred Term, Safety Population.

|  |  | **Patient Count (%) by Treatment** |  |
| --- | --- | --- | --- |
| **System Organ Class**<br>**Preferred Term** | **Total**<br>**(N=334)** | **FPV**<br>**(N=168)** | **Placebo**<br>**(N=166)** |
| Any Related Adverse Event | 62 (19%) | 35 (21%) | 27 (16%) |
| Investigations | 19 (6%) | 9 (5%) | 10 (6%) |
| *Alanine aminotransferase increased* | 1 (0%) | 1 (1%) | 0 (0%) |
| *Blood creatinine increased* | 1 (0%) | 0 (0%) | 1 (1%) |
| *Blood triglycerides increased* | 1 (0%) | 0 (0%) | 1 (1%) |
| *Blood uric acid increased* | 2 (1%) | 1 (1%) | 1 (1%) |
| *Haemoglobin decreased* | 1 (0%) | 1 (1%) | 0 (0%) |
| *Hepatic enzyme increased* | 11 (3%) | 6 (4%) | 5 (3%) |
| *Lipids abnormal* | 1 (0%) | 0 (0%) | 1 (1%) |
| *Liver function test increased* | 1 (0%) | 0 (0%) | 1 (1%) |
| Metabolism and nutrition disorders | 10 (3%) | 9 (5%) | 1 (1%) |
| *Gout* | 1 (0%) | 1 (1%) | 0 (0%) |
| *Hyperuricaemia* | 9 (3%) | 8 (5%) | 1 (1%) |
| Respiratory, thoracic and mediastinal disorders | 25 (7%) | 14 (8%) | 11 (7%) |
| *Acute respiratory distress syndrome* | 13 (4%) | 7 (4%) | 6 (4%) |
| *Acute respiratory failure* | 1 (0%) | 0 (0%) | 1 (1%) |
| *Hypoxia* | 1 (0%) | 1 (1%) | 0 (0%) |
| *Respiratory failure* | 11 (3%) | 7 (4%) | 4 (2%) |
| N = Total number of patients in the specified treatment group |  |  |  |
| Percentages are based on the total number of patient's in the specified treatment group under the Safety Analysis set |  |  |  |

N = Total number of patients in the specified treatment group
Percentages are based on the total number of patient's in the specified treatment group under the Safety Analysis set

During Stage I of the study, 33 (10%) patients experienced at least one SAE, with 19 (11%) patients in the FPV and 14 (8%) in the placebo group. Maximum number of SAEs reported were associated with respiratory, thoracic, and mediastinal disorders, including acute respiratory distress syndrome in 13 (4%) patients [7 (4%) patients in FPV and 6 (4%) in placebo group] and acute respiratory failure/respiratory failure in 12 (4%) patients [7 (4%) patients in FPV and 5 (3%) patients in placebo group].

Overall, 2 (1%) patients experienced at least one TEAE in Stage II with, 1 (1%) patient each in the FPV and placebo groups. SAE in the FPV group was attributed to hepatobiliary disorders (chronic calculous cholecystitis) and was assessed as not related to FPV treatment.

## Discussion

The demand for a reliable, safe, and effective anti-viral treatment option for COVID-19 is largely unmet and continues to drive clinical research efforts. The current Phase 3 study did not find FPV to be effective in comparison to placebo in moderate to severe COVID-19 patients. Nevertheless, some trends in favour of FPV were observed on *post-hoc* subgroup analysis, in the patients with ‘lower’ baseline NEWS-2 based clinical risk categories of ‘low’ and ‘low-medium’, for the primary efficacy endpoint ‘time to resolution of hypoxia’ and also for some of the key secondary endpoints, such as time to hospital discharge, and improvement in clinical status score by 1-point over baseline on the WHO 10-point ordinal scale. These trends observed in the lower clinical risk categories suggest that patients with lower COVID-19 disease severity at start of treatment may benefit from FPV treatment. A corollary inference could be that antiviral drugs should be initiated early in the course of infection to provide optimal benefit to patients. Currently, a placebo-controlled, phase 3 PRESECO (PREventing SEvere COvid-19) trial is ongoing to evaluate the efficacy and safety of FPV when initiated earlier in the course of COVID-19 in the outpatient setting[15].

The risk of rapid COVID-19 progression underlines the need for earlier intervention with an effective antiviral treatment in COVID-19[16]. In a prospective, randomized, open-label trial of early versus late FPV in hospitalized patients with COVID-19 conducted at 25 hospitals across Japan, a trend toward better viral clearance on day 6 was seen in the early treatment group as compared to late treatment group [66.7% versus 56.1%, adjusted hazard ratio (aHR) (95% CI): 1.42 (0.76–2.62)]. In line with this trend, faster defervescence was reported in the early treatment group [2.1 days versus 3.2 days, aHR (95% CI): 1.88 (0.81–4.35 (p = 0.048)][17]. In another retrospective study published by Yu T, *et al*., (2020), early antiviral treatment demonstrated 7 days shorter virus clearance time when compared with late antiviral treatment. After virus clearance, the group with early antiviral treatment showed milder illness than the group with late antiviral treatment. The study concluded that early antiviral treatment could effectively shorten the virus clearance time and prevent the rapid progression of COVID-19[18].

Conflicting findings have been reported in the current literature regarding the efficacy of FPV in the treatment of COVID-19. In an observational study of Favipiravir, in a total of 10,986 SARS-CoV-2 infected patients from 765 hospitals, clinical improvement at 7 and 14 days was 72.6 % and 86.5%, 63.4% and 77.2%, and 46.6% and 60.4% for mild, moderate, and severe diseases, respectively. The mortality rates within a month from hospitalization were 3.6%, 13.2%, and 27.6% for mild, moderate, and severe diseases. Both the clinical course and outcomes were poorer in older and severe patients[11]. Clinical trials conducted in COVID-19 patients where FPV treatment was compared to placebo, hydroxychloroquine, lopinavir/ritonavir, umifenovir (arbidol) or standard of care showed clinically significant trends (decrease in the duration of hospital stay and the need for mechanical ventilation) on a number of clinically pertinent endpoints[17,19–22].

In a randomized, open-label, parallel-arm, multicenter, Phase 3 study by Udwadia *et al*., (2021) it was suggested that despite failure to achieve statistical significance on the primary endpoint of time to RT-PCR negativity, early administration of oral FPV may reduce the duration of clinical signs and symptoms in patients with mild-to-moderate COVID-19, as demonstrated by the significantly decreased time to clinical cure[22]. The current study also demonstrated favourable trends on early administration of oral FPV in patients with ‘lower’ NEWS-2 clinical risk category.

The adverse events reported with FPV in the current study were consistent with previous experience with the drug. There were no unexpected adverse events in the study. The fatalities reported in the study were likely attributable to progression and complications associated with COVID-19 in both groups.

In the present study, higher blood uric acid levels were reported at days 10 and 28 or upon discharge in FPV group compared to placebo group patients. In the FPV group, 5.4% reported hyperuricaemia, of which 5% were considered treatment related in the current study. This was much lower compared to the incidence of 15.5% reported in trials conducted in Japan[23] and 13.8% in Russia and China[24]. This suggests that Asian population could be more susceptible to the FPV-associated hyperuricaemia but the reasons for this remain unknown. One possibility to consider for these variations across demographics could be related to variations in the enzymatic activity of aldehyde oxidase and xanthine oxidase - the key enzymes responsible for metabolizing FPV. Functionally inactive human aldehyde oxidase (hAOX1) allelic variants as well as variants encoding enzymes with different catalytic activities exist within the human population. Future investigations into these allelic variants could offer further insight into these differences[25]. To date, no evidence has emerged that FPV-associated hyperuricaemia is associated with clinical manifestations. Longer trial follow-up periods would be required to assess this risk fully[26].

Further studies evaluating the role of FPV in treating the disease in specific population groups and disease severity categories are warranted. The study analysis faced challenges from missing data at various timepoints due to large number of discontinuations and early discharges. Despite the limitation, the present study showed trends suggesting that FPV is associated with better clinical outcomes in some subgroups of patients (younger age, lower BMI, and lower baseline clinical risk based on NEWS-2) with tolerability and safety of FPV being comparable to placebo.

## Conclusion

The trial did not find favipiravir to be effective in moderate to severe COVID-19 patients but trends in favour of favipiravir were observed in the NEWS-2 low to low-medium clinical risk subgroup for several clinically meaningful endpoints. No new or unexpected adverse events were noted. Future studies should evaluate the efficacy of the drug when administered early in the disease.

## Data Availability

All data produced in the present study are available upon reasonable request to the authors

## Acknowledgements

The authors would like to thank all members of the Kuwait Clinical Trial Group*, DM Rao and team from Dr. Reddy’s (Regulatory support), David He (Biostatistician), Dr Jyoti Rao (Medical writer) and Navitas Life Sciences without whom the conduct of study would not be possible.

## Conflict of Interest

Drs. Srinivas Shenoy and Sagar Munjal are paid employees of Dr. Reddy’s Laboratories and didn’t receive any additional compensation for this study.

Dr Salman Al-Sabah was the Principal Investigator and National Co-Ordinator (Kuwait) for the trial, while Drs. Sarah Al Youha, Mohammad Alghounaim, Sulaiman Almazeedi and Yousef Alshamali were sub-Investigators. The Investigators and sub-investigators are employees of various public sector hospitals in the State of Kuwait and did not receive any financial compensation for this study.

Dr Richard H Kaczynski is a consultant to Fujifilm Toyoma Chemical Co. Ltd. and Chief Medical Officer of AiPharma. He contributed significantly to study design and did not receive any additional compensation for this study.

## Notes

### Competing Interest Statement

Drs. Srinivas Shenoy and Sagar Munjal are paid employees of Dr. Reddy s Laboratories and did not receive any additional compensation for this study.

### Clinical Trial

NCT04529499
IND148286

### Funding Statement

The study was funded by Dr. Reddys Laboratories Ltd. and Global Response Aid - a subsidiary of Agility Inc.

### Author Declarations

Ministry of Health Kuwait Ethics committee gave ethical approval for 3 study centres in Kuwait Abu Dhabi - IRB Committee, The Department of Health (DoH), Abu Dhabi approval was obtained for the UAE site

