## Supplementary Data for "Favipiravir In Adults with Moderate to Severe COVID-19: A Phase 3 Multicentre, Randomized, Double-Blinded, Placebo-Controlled Trial"

**Supplementary Tables and Data**

Table 1: Details of Kuwait Clinical Trial group

| **S.No** | **Investigator Name** | **Affiliation** | **Contribution** |
| --- | --- | --- | --- |
| 1 | Mohammad Al Saffar | Jaber Al Ahmad Hospital | Co-investigator, clinical lead, supervision |
| 2 | Mariam Boulbanat | Jaber Al Ahmad Hospital | Co-investigator, clinical lead, supervision |
| 3 | Mohammad Al Humaidan | Kuwait Field Hospital-Mishref | Principal investigator, supervision |
| 4 | Naela Al Mazeedi | Farwaniya Hospital | Principal investigator, supervision |
| 5 | Ali Al Harbi | Jaber Al Ahmad Hospital | Co-investigator, lead pharmacist, supervision, project administration |
| 6 | Buthaina Alkandari | Jaber Al Ahmad Hospital | Co-investigator, lead radiologist, supervision |
| 7 | Fahad Al Abdulghani | Al Amiri Hospital | Co-investigator, radiological interpretation |
| 8 | Abdullatif Al Busairi | Jaber Al Ahmad Armed Forces Hospital | Co-investigator, radiological interpretation |
| 9 | Salem Al Qahtani | Farwaniya Hospital | Co-investigator, clinical lead, supervision |
| 10 | Wassim Chehadeh | Kuwait University | Co-investigator, lab data analysis, supervision |
| 11 | Kelly Schrapp | Jaber Al Ahmad Hospital | Co-investigator, ICU lead, conceptualization |

Table 2: Criteria applied for Randomization Severity Strata Categorization

| **Severity Strata** | **Criteria*** |
| --- | --- |
| Moderate | Patients clinically assigned ‘moderate’ [symptoms of moderate illness with COVID-19, which could include any symptom of fever, cough, sore throat, malaise, headache, muscle pain, gastrointestinal symptoms or shortness of breath with exertion and clinical signs suggestive of moderate illness with COVID-19, such as respiratory rate ≥20 breaths per minute or saturation of oxygen (SpO_2_) >93% on room air at sea level or heart rate ≥90 beats per minute] |
| Severe | Patients clinically assigned ‘severe’ (symptoms suggestive of severe systemic illness with COVID-19, which could include any symptom of moderate illness or shortness of breath at rest, or respiratory distress and clinical signs indicative of severe systemic illness with COVID-19, such as respiratory rate ≥30 per minute or heart rate ≥125 per minute or SpO_2_ ≤93% on room air at sea level or PaO_2_/FiO_2_ <300) |
| *Note: The severity is defined as per the FDA Guidance document COVID-19: Developing Drugs and Biological Products for Treatment or Prevention - Guidance for Industry Final Document dated May 2020 | |

Table 3: National Early Warning Score-2 (NEWS-2) Scoring Scale

| **Variable** | | **Points** |
| --- | --- | --- |
| **Respiratory rate, breaths per minute** | ≤8 | 3 |
|  | 9-11 | 1 |
|  | 12-20 | 0 |
|  | 21-24 | 2 |
|  | ≥25 | 3 |
| **SpO_2_ (on room air or supplemental)** | ≤91% | 3 |
|  | 92-93% | 2 |
|  | 94-95% | 1 |
|  | ≥96% | 0 |
| **SpO_2_ (if patient has hypercapnic respiratory failure)** | ≤83% | 3 |
|  | 84-85% | 2 |
|  | 86-87% | 1 |
|  | 88-92%, ≥93% on room air | 0 |
|  | 93-94% on supplemental oxygen | 1 |
|  | 95-96% on supplemental oxygen | 2 |
|  | ≥97% on supplemental oxygen | 3 |
| **Room air or supplemental oxygen** | Supplemental oxygen | 2 |
|  | Room air | 0 |
| **Temperature** | ≤35.0°C (95°F) | 3 |
|  | 35.1-36.0°C (95.1-96.8°F) | 1 |
|  | 36.1-38.0°C (96.9-100.4°F) | 0 |
|  | 38.1-39.0°C (100.5-102.2°F) | 1 |
|  | ≥39.1°C (102.3°F) | 2 |
| **Systolic BP, mmHg** | ≤90 | 3 |
|  | 91-100 | 2 |
|  | 101-110 | 1 |
|  | 111-219 | 0 |
|  | ≥220 | 3 |
| **Pulse, beats per minute** | ≤40 | 3 |
|  | 41-50 | 1 |
|  | 51-90 | 0 |
|  | 91-110 | 1 |
|  | 111-130 | 2 |
|  | ≥131 | 3 |
| **Consciousness** | Alert | 0 |
|  | New-onset confusion (or disorientation/agitation), responds to voice, responds to pain, or unresponsive | 3 |

Note: *Interpretation of NEWS-2 score:*

| **NEWS-2 Score** | **Clinical risk** | **Frequency of monitoring** | **Response** |
| --- | --- | --- | --- |
| **0-4** | Low | Minimum every 12 hrs if score of 0  Minimum every 4-6 hrs if score 1-4 | Assessment by a competent registered nurse or equivalent, to decide change in frequency of clinical monitoring or escalation of care |
| **Score of 3 in any individual parameter** | Low-medium | Minimum every hour | Urgent review by a ward-based doctor, to decide change in frequency of clinical monitoring or escalation of care |
| **5-6** | Medium |  | Urgent review by a ward-based doctor or acute team nurse, to decide if critical care team assessment is needed |
| **≥7** | High | Continuous monitoring of vital signs | Emergent assessment by a clinical team or critical care team and usually transfer to higher level of care |

Table 4: Summary of Subgroup Analysis of Time to Event Endpoints from Baseline by Age*

|  | **Age < 50 years** | | **Age ≥ 50 years** | |
| --- | --- | --- | --- | --- |
| **Parameters/Variables** | **FPV**  **(N=70)** | **Placebo**  **(N=74)** | **FPV**  **(N=105)** | **Placebo**  **(N=104)** |
| ***Time to Resolution of Hypoxia (Primary endpoint)*** | | | | |
| No of Patients at clinical risk** | 61 | 61 | 96 | 97 |
| No of Events [Time frame: Up to 28 days]  (% Reached Resolution Endpoint) | 51 (83.6%) | 46 (75.4%) | 71 (74.0%) | 72 (74.2%) |
| Time to event, median days | 7 | 6 | 7 | 9 |
| Hazard Ratio (95% CI) | 0.95 (0.639, 1.419) | - | 1.1 (0.789, 1.522) | - |
| *P-*value^∮^ | 0.81 |  | 0.59 |  |
| ***Time to Hospital Discharge by Treatment*** | | | | |
| No of Events (% Reached Discharge Endpoint) | 65 (93.0%) | 66 (89.2%) | 92 (87.6%) | 90 (86.5%) |
| Time to event, median days | 9 | 10 | 11 | 11 |
| Hazard Ratio (95% CI) | 1.06 (0.754, 1.497) | - | 1.11 (0.832, 1.490) | - |
| *P-*value^∮^ | 0.73 | - | 0.47 | - |
| ***Time to improvement by 1-point from baseline in WHO 10-point clinical status score*** | | | | |
| No of Events (% Reached Improve 1 Point Endpoint) | 56 (80.0%) | 55 (74.3%) | 73 (69.5%) | 74 (71.2%) |
| Time to event, median days | 7 | 6 | 7 | 9 |
| Hazard Ratio (95% CI) | 0.99 (0.679, 1.431) | - | 1.07 (0.773, 1.477) | **-** |
| *P-*value^∮^ | 0.94 |  | 0.69 |  |
| CI: Confidence Interval  * Age group is presented into 2 categories: < 50 years and ≥ 50 years  ****** Patients with Baseline WHO 10-Point Clinical Status Score > 4 (Hospitalized: no oxygen therapy) (N=315) were included in the Primary endpoint analysis (Time to Resolution of Hypoxia); 38 Patients with score of 4 were not included.  ^∮^ From Cox proportional hazards model | | | | |

Table 5: Summary of Subgroup Analysis of Time to Event Endpoints from Baseline by BMI*

|  | **BMI < 30 Kg/m^2^** | | **BMI ≥ 30 Kg/m^2^** | |
| --- | --- | --- | --- | --- |
| **Parameters/Variables** | **FPV**  **(N=85)** | **Placebo**  **(N=86)** | **FPV**  **(N=84)** | **Placebo**  **(N=86)** |
| ***Time to Hospital Discharge by Treatment*** | | | | |
| No of Events (% Reached Discharge Endpoint) | 75 (88.2%) | 77 (89.5%) | 76 (90.5%) | 75 (87.2%) |
| Time to event, median days | 10 | 10 | 11 | 11 |
| Hazard Ratio (95% CI) | 0.96 (0.700, 1.324) | - | 1.22 (0.887, 1.684) | - |
| *P-*value^∮^ | 0.82 | - | 0.22 | - |
| ***Time to improvement by 1-point from baseline in WHO 10-point clinical status score*** | | | | |
| No of Events (% Reached Improve 1 Point Endpoint) | 58 (68.2%) | 68 (79.1%) | 66 (78.6%) | 59 (68.6%) |
| Time to event, median days | 7 | 6 | 7 | 9 |
| Hazard Ratio (95% CI) | 0.74 (0.521, 1.054) | - | 1.41 (0.993, 2.007) | **-** |
| *P-*value^∮^ | 0.095 |  | 0.055 |  |
| BMI: Basal Metabolic Index; CI: Confidence Interval  * BMI data is presented into 2 categories: < 30 and ≥ 30 Kg/m^2^  ** The post-hoc exploratory main effects model did not show significant association between covariate BMI and time to resolution of hypoxia endpoint .  ^∮^ From Cox proportional hazards model | | | | |

Table 6: Proportion of Patients with WHO 10-Point Clinical Status Score Improving at Least 1-Point from Baseline by Treatment and Day, ITT Population (N=353)

|  | | | **By Treatment Group** | |
| --- | --- | --- | --- | --- |
| **Visit** | **Statistics/ Response Category** | **Total (N=353)** | **Favipiravir (N=175)** | **Placebo (N=178)** |
| Day 4 | Yes | 73 (20.7%) | 42 (24.0%) | 31 (17.4%) |
|  | Data Missing | 30 (8.5%) | 12 (6.9%) | 18 (10.1%) |
| Day 7 | Yes | 122 (34.6%) | 62 (35.4%) | 60 (33.7%) |
|  | Data Missing | 78 (22.1%) | 40 (22.9%) | 38 (21.3%) |
| Day 10 | Yes | 97 (27.5%) | 47 (26.9%) | 50 (28.1%) |
|  | Data Missing | 160 (45.3%) | 83 (47.4%) | 77 (43.3%) |
| Day 14 | Yes | 18 (5.1%) | 8 (4.6%) | 10 (5.6%) |
|  | Data Missing | 290 (82.2%) | 144 (82.3%) | 146 (82.0%) |
| Day 21 | Yes | 4 (1.1%) | 2 (1.1%) | 2 (1.1%) |
|  | Data Missing | 328 (92.9%) | 161 (92.0%) | 167 (93.8%) |
| Day 28 or Discharge | Yes | 223 (63.2%) | 118 (67.4%) | 105 (59.0%) |
|  | Data Missing | 29 (8.2%) | 10 (5.7%) | 19 (10.7%) |
| Cross-sectional data, not cumulative.  No statistical inference was performed due to missing data associated with key efficacy endpoints such as resolution of hypoxia and hospital discharge. | | | | |

Table 7: Proportion of Patients with WHO 10-Point Clinical Status Score Improving at Least 2-Point from Baseline by Treatment and Day, ITT Population (N=353)

|  | | | **By Treatment Group** | |
| --- | --- | --- | --- | --- |
| **Visit** | **Statistics/ Response Category** | **Total (N=353)** | **Favipiravir (N=175)** | **Placebo (N=178)** |
| Day 4 | Yes | 4 (1.1%) | 3 (1.7%) | 1 (0.6%) |
|  | Data Missing | 30 (8.5%) | 12 (6.9%) | 18 (10.1%) |
| Day 7 | Yes | 8 (2.3%) | 4 (2.3%) | 4 (2.2%) |
|  | Data Missing | 78 (22.1%) | 40 (22.9%) | 38 (21.3%) |
| Day 10 | Yes | 15 (4.2%) | 7 (4.0%) | 8 (4.5%) |
|  | Data Missing | 160 (45.3%) | 83 (47.4%) | 77 (43.3%) |
| Day 14 | Yes | 3 (0.8%) | 0 (0.0%) | 3 (1.7%) |
|  | Data Missing | 290 (82.2%) | 144 (82.3%) | 146 (82.0%) |
| Day 21 | Yes | 0 (0%) | 0 (0%) | 0 (0%) |
|  | Data Missing | 328 (92.9%) | 161 (92.0%) | 167 (93.8%) |
| Day 28 or Discharge | Yes | 31 (8.8%) | 15 (8.6%) | 16 (9.0%) |
|  | Data Missing | 29 (8.2%) | 10 (5.7%) | 19 (10.7%) |
| Cross-sectional data, not cumulative.  No statistical inference was performed due to missing data associated with key efficacy endpoints such as resolution of hypoxia and hospital discharge. | | | | |
